# Cost of In-patient Management of Covid-19 Patients in a Tertiary Hospital in Kuwait

**DOI:** 10.1101/2022.11.21.22282601

**Authors:** Amrizal Muhammad Nur, Syed Mohamed Aljunid, Mohammad Almari

## Abstract

**Background:** Among the GCC countries affected by COVID-19 infections, Kuwait was impacted with 658,520 cases and 2,563 deaths as reported by WHO on September 30, 2022. However, the impact of the COVID-19 epidemic on the economy of Kuwait especially in health sector is unknown.

**Objective:** The aim of this study is to determine the total cost of COVID-19 in-patient management in Kuwait.

**Method:** Retrospective design was employed in this study. A total 485 Covid-19 patients admitted to a tertiary hospital assigned to manage Covid-19 cases was randomly selected for this study from 1st May to 31st September 2021. Data on sociodemographic, length of stay (LOS), discharge status and comorbidity were obtained from the patients’ medical records. Among others, data on cost in this study cover administration, utility, pharmacy, radiology, laboratory, nursing, and ICU costs. The unit cost per admission was imputed using a step-down costing method with three levels of cost centers. The unit cost was multiplied by the individual patient’s length of stay to obtain the cost of care per patient per admission.

**Findings:** The mean cost of Covid-19 inpatient per episode of care was KD 2,216 (SD=2,018) equals to US$ 7,344 (SD=6,688) with the average length of stay of 9.4 (SD=8.5) days per admission. The total treatment costs of Covid-19 inpatient (n=485) were estimated to be KD 1,074,644 (US$ 3,561,585), in which the physician and nursing care cost were the largest share of costs (42.1%) with KD 452,154 (US$ 1,498,529). The second- and third-largest costs were intensive care (20.6%) of KD 221,439 (US$ 733,893) and laboratory costs (10.2%) of KD 109,264 (US$ 362,123). The average cost for severe Covid-19 patient was KD 4,626 (US$ 15,332), which is almost three times higher than the non-severe patients of KD 1,544 (US$ 5,117).

**Conclusion:** The cost of managing Covid-19 cases is substantial. The cost information can assist hospital managers and policymakers in designing more efficient interventions, especially for the management of high-risk groups.

## Introduction

Initial cases of novel coronavirus (2019-nCoV)-infected pneumonia (NCIP) occurred in Wuhan, Hubei Province, China, in December 2019 and January 2020 [1]. This virus has caused widespread pandemics with high morbidity and mortality [2]. Globally, there have been 614,385,693 confirmed cases of COVID-19, including 6,522,600 deaths and 325,602 new cases reported by the WHO. As of September 27, 2022, 12,677,499,928 vaccine doses were administered [at 3]. Among the GCC countries affected by COVID-19 infections, Kuwait was impacted, with 658,520 cases and 2,563 deaths A total of 8,214,656 vaccine doses have been administered in Kuwait as reported by WHO on September 30, 2022, [4].

Many countries closed their borders, stopped all flights, and had to reduce their affairs with other countries. These measures caused significant economic shrinkage worldwide, with businesses closing, increasing unemployment, rising inflation, and the interruption of production and shipping [5]. The impact of COVID-19 on the global economy is more severe because these infections reduce labor supply. Quarantines, regional lockdowns, and social distancing have been adopted to contain the virus. Workplace closures disrupt supply chains and lower productivity. Layoffs, income declines, fear of contagion, and heightened uncertainty make people spend less, triggering further business closures and job loss. All these lead to the shutdown of a significant portion of the economy (IMF) [6].

World leaders have already been warned of the sustainability of health care systems before the current pandemic hit [7, 8]. The rapid and global spread of the pandemic has exacerbated existing problems and created new issues that have challenged decision-makers and negatively impacted the health of the populace [9]. To mitigate the impact of this and future pandemics, decision makers need to understand how their health systems are impacted. Of particular importance is understanding healthcare resource use (e.g., length of hospital stays) and subsequent costs of the pandemic [10]. Decision makers can use this knowledge to plan resources and allocate budgets for future health crises. Many people have lost their jobs due to the pandemic. Many economic and social sectors have been indirectly affected by Covid-19. Some of the business sectors that the pandemic has already impacted include general transport, general insurance, manufacturing, and some forms of healthcare [11]. The results of a study in the United States indicated that approximately 50% of participants reported income and wealth losses due to the coronavirus, with average losses of US$ 5,293 and US$ 33,482, respectively [12]. A study in Russia showed that the socioeconomic burden of COVID-19 would amount to 4.6 trillion rubles (US$ 71.1 billion) or 4% of GDP [13]. The prevention and treatment of COVID-19 can impose a considerable economic burden on many people and societies. According to previous studies [14], all confirmed cases of Covid-19 should receive inpatient care. Furthermore, Covid-19 patients often require costly treatment, such as mechanical ventilation and extracorporeal membrane oxygenation, potentially considerably increasing healthcare costs [15]. A study in the United States (2020) indicated that a single symptomatic Covid-19 infection accounted for a median direct medical cost of US$ 3, 045. They also reported that Covid-19 coronavirus in the United States of America could result in direct medical costs incurred during the infection from US$ 163.3 billion if 20% of the population infected to US$ 654.0 billion if 80% of the population is infected [16].In a study in China, the total estimated healthcare and societal costs associated with Covid-19 were US$ 0.62 billion and US$ 383.02 million, respectively [17]. Kuwait is a country located in the Middle East with a population of 4,328,553 million (2021). The GDP per capita was estimated at US$ 24,811. The current health expenditure is 5.5% of the GDP [27,28].

The estimation of coronavirus disease burden on the health system can not only assist policymakers in effectively allocating resources and prioritizing various measures, but also emphasizes the importance of long-term planning for sustainable financing of similar conditions in the future. To the best of our knowledge, the impact of the COVID-19 pandemic on the economy of Kuwait, especially in the health sector, is unknown. This is the first study on health economic evaluation in Kuwait. This study aimed to determine the total cost of COVID-19 inpatient management in a tertiary government hospital in Kuwait.

## Methods

This study was a partial economic evaluation with a cross-sectional retrospective design, and the management cost of inpatients admitted with COVID-19 and positive PCR test results in a tertiary hospital in Kuwait between 1st May to 31st September 31, 2021. Patient clinical data were obtained from 485 randomly selected medical records (MR) of patients and extracted from electronic patient records through the Hospital Information System (HIS) using patient data collection tool (appendix 1). Clinical data included sociodemographic characteristics (age, sex, etc.), length of stay (LOS), discharge status, primary diagnosis, comorbidity, medical or surgical procedures, laboratory tests, radiology tests, and physiotherapy services. The full list of the information collected in this study is given in an appendix 1. The management cost of Covid-19 patient was carried out imputed using a step-down costing method, which was calculated from the provider ‘s perspective. Hospital costing data were collected using the costing data collection tool in an excel sheet format that organizes costing data into three levels of cost centers: overhead cost centers (e.g., administration, consumables, maintenance, etc.), intermediate cost centers (e.g., pharmacy, radiology, etc.), and final cost centers, all wards and clinics (appendix 2). To estimate the cost for each cost center, both capital costs (building, equipment, and furniture costs) and recurrent costs (staff salary and other operational costs except salary) were combined. Information on activities that reflect the workload, such as the number of discharges, inpatient days, number of visits, and floor space, are gathered from medical records and engineering departments to determine appropriate allocation factors.

### Computing of the provider cost

There are seven steps in computing the unit cost using the stepdown costing approach:1. Defining the cost centers, and 2.Grouped cost centers into (overhead cost centers, intermediate cost centers, and final/patient care cost centers); 3. Obtain the total cost of each overhead, intermediate, and final care cost center, 4. Decide on units to distribute costs; and 5. Allocate the costs of overhead cost centers to intermediate and final care cost centers; 6. Allocate the costs of intermediate-to-final care cost centers; Relate the unit cost to the individual patient’s length of stay to obtain the cost of care per patient per discharge. These 7 steps were adopted and modified method from WHO, Shepard, DS et.al (2000) and it has been used for costing research method many times by Amrizal et.al [19,20]. The study was approved by the Research and Ethics Committee of Kuwait Ministry of Health (Approval Code:1502/2020). The study does not require informed consent because the data was extracted from Jaber Al Ahmad hospital information system database and did not involve interviews of patients.

## Results

### Patient Demographics and other characteristics

A total of 485 hospitalized patients with Covid-19 were included in this study. The mean and SD age of patients was estimated at 52.9 (SD=18.1 years) and median ages 55 years. Most of the patients were Kuwaitis patients (87.0%), females (54.6%), admitted to the general ward (79.6%) and ICU (20.4%), discharged home, or recovered 73.4%. Most of the patients were symptomatic (94.8%). The mean length of hospital stay was 9.4 ± 8.5 days. Covid-19 disease types were mild 139 (28.5%), moderate 259 (53.1%), and severe 87 (18.4%) (Table-1).

**Table-1:** Demographics and other characteristics of Covid-19 Patients.

| **Patient Demographic and Characteristics** | **Mild**<br>**(N = 139)** | **Moderate**<br>**(N = 259)** | **Severe**<br>**(N = 87)** | **Overall**<br>**(N = 485)** |
| --- | --- | --- | --- | --- |
| **Gender** |  |  |  |  |
| Female | 78 (56.1%) | 146 (56.4%) | 41 (47.1%) | 265 (54.6%) |
| Male | 61 (43.9%) | 113 (43.6%) | 46 (52.9%) | 220 (45.4%) |
| **Age (years)** |  |  |  |  |
| Mean ± SD | 47.2 ± 19.1 | 53.7 ± 17.6 | 59.9 ± 14.9 | 52.9 ± 18.1 |
| Median [Min-Max] | 50 (0-89) | 55 (0-97) | 60 [15-94] | 55 [0-97] |
| **LOS (days)** |  |  |  |  |
| Mean ± SD | 4.5 ± 3.4 | 8.6 ± 5.2 | 19.6 ± 12.8 | 9.4 ± 8.5 |
| Median [Min-Max] | 4 [1-29] | 7 [1-43] | 17 [2-57] | 7 [1-57] |
| **Nationality** |  |  |  |  |
| Kuwaiti | 130 (93.5%) | 229 (88.4%) | 63 (72.4%) | 422 (87.0%) |
| Non-Kuwaiti | 9 (6.5%) | 30 (11.6%) | 24 (27.6%) | 63 (13.0%) |
| **Admission** |  |  |  |  |
| General ward | 138 (99.3%) | 245 (94.6%) | 3 (3.4%) | 386 (79.6%) |
| ICU | 1 (0.7%) | 14 (5.4%) | 84 (96.6%) | 99 (20.4%) |
| **Disease Type** |  |  |  |  |
| Asymptomatic | 8 (5.8%) | 7 (2.7%) | 10 (11.5%) | 25 (5.2%) |
| Symptomatic | 131 (94.2%) | 252 (97.3%) | 77 (88.5%) | 460 (94.8%) |
| **Discharge Status** |  |  |  |  |
| Recovered | 121 (87.1%) | 223 (86.1%) | 12 (13.8%) | 356 (73.4%) |
| Transfer to other hospital | 1 (0.7%) | 9 (3.5%) | 21 (24.1%) | 31 (6.4%) |
| DAMA | 17 (12.2%) | 25 (9.7%) | 0 (0.0%) | 42 (8.7%) |
| Died | 0 (0.0%) | 2 (0.8%) | 54 (62.1%) | 56 (11.5%) |
| Total | 139 (28.7%) | 259 (53.4%) | 87 (17.9%) | 485 (100%) |

As shown in (Table 2), the Treatment cost per admission and ALOS of mild condition: KD 1064 (SD=795) equals US$ 3,526 (SD=2,635) and ALOS 4.5 days (SD=3.4). The treatment cost per admission and ALOS of moderate condition is KD 2,024 (SD=1,233), equal to US$ 6,708 (SD=4,086) and ALOS 8.6 days (SD=5.2), respectively. The treatment cost per admission and ALOS of severe condition is KD 4,626 (SD=3,035) equal to US$ 15,331 (SD=10,059) and ALOS 19.6 days (SD=12.8), respectively. The overall treatment cost per patient per admission was KD 2,216 (SD=2,018), equal to US$ 7,344 (SD=6,688), with an average Length of Stay (ALOS) of 9.4 days (SD=8.5). In this study, US dollars were used for international comparison. Average exchange rate in 2021 (1 KD= US$ 3.3142).

**Table 2:** Mean Cost and Length of Stay (LOS) of Covid-19 Patients by Severity.

| Types of Severity | N | Mean LOS<br>(SD) (days) | Mean<br>Cost (KD) | SD<br>(KD) | Mean Cost<br>(US\$) | SD<br>(US\$) |
| --- | --- | --- | --- | --- | --- | --- |
| Mild | 139 | 4.5 (SD=3.4) | 1,064 | 795 | 3,526 | 2,635 |
| Moderate | 259 | 8.6 (SD=5.2) | 2,024 | 1,233 | 6,708 | 4,086 |
| Severe | 87 | 19.6 (SD=12.8) | 4,626 | 3,035 | 15,331 | 10,059 |
| Averages | 485 | 9.4 (SD=8.5) | 2,216 | 2,018 | 7,344 | 6,688 |

Notes: SD = Standard Deviation KD = Kuwaiti Dinar US\$ = US Dollar Average exchange rate in year 2021 (1 KD= 3.3142 US\$).

As shown in (Table 3), The treatment cost per admission and ALOS of patients in the general ward: KD 1,670 (SD=1,175) equals US$ 5,535 (SD=3,894) and ALOS 7.1 days (SD=4.9). The treatment cost per admission and ALOS of patients in the ICU were as follows: KD 4,342 (SD=2,992) equals US$ 14,390 (SD=9,916) and ALOS 18.4 days (SD=12.7). In general, the treatment cost per patient per admission in averages: KD 2,216 (SD=2,018) equals US$ 7,344 (SD=6,688) and the average Length of Stay (ALOS) per patient per admission on averages: 9.4 days (SD=8.5).

**Table 3:**
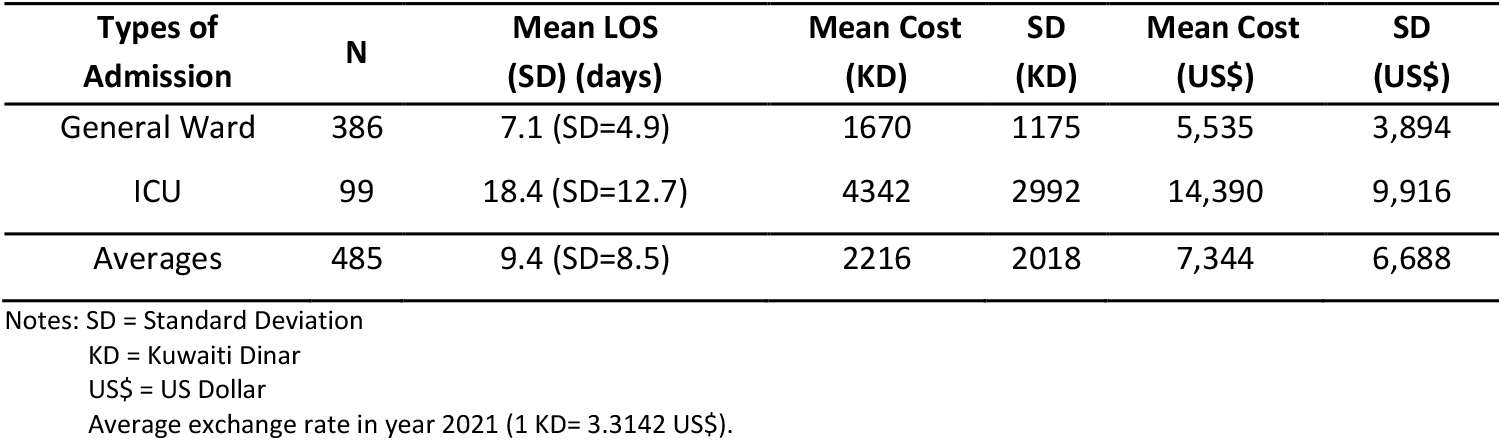
Mean Cost and Length of Stay (LOS) of Covid-19 Patients by General Ward and ICU.

Regarding the total treatment costs and the percentages of cost component for 485 covid-19 inpatients, there were estimated to be KD 1,074,644 (US$ 3,561,585), in which the physician and nursing care cost were the largest share of costs (42.1%) with KD 452,154 equals to US$ 1,498,529. Following that, the main costs were intensive care costs (20.6%) with KD 221,439 equal to US$ 733,893, laboratory costs (10.2%) KD 109,264 equal to US$ 362,123, administration costs (6.7%) KD 71,479 equal to US$ 236,896 and drug costs (5.5%) KD 58,611 equals to US$ 194,249. The average treatment cost for Covid-19 patient with severe conditions was almost three times higher than that for non-severe patients. The average cost of severe patients was KD 4,626 (SD=3,035), equal to US$ 15,332 (SD=10,059), and non-severe patient were KD 1,544 (SD=1,014), equal to US$ 5,117 (SD=3,361). In general, the average treatment cost per patient per admission, KD 2,216 (SD=2,018), equals US$ 7,344 (SD=6,688) (Table 4 and Figure 1).

**Table 4:** Total Cost of Each Cost Component by Severe and Non-Severe of Covid-19 Patients.

| Cost Component | % | Total Cost All Patient (n=485) (KD/US\$) | Mean Cost per Patient (KD/US\$)±(SD) | SD | Mean Cost per Non-Severe Patient (KD/US\$)±(SD) | SD | Mean Cost per Severe Patient (KD/US\$)±(SD) | SD |
| --- | --- | --- | --- | --- | --- | --- | --- | --- |
| Administration | 6.7% | 71,479 (KD) | 147 (KD) | 134 (KD) | 103 (KD) | 68 (KD) | 308 (KD) | 202 (KD) |
| | | 236,896 (US\$) | 487 (US\$) | 444 (US\$) | 341 (US\$) | 225 (US\$) | 1,021 (US\$) | 669 (US\$) |
| Maintenance | 0.9% | 10,094 (KD) | 21 (KD) | 19 (KD) | 15 (KD) | 10 (KD) | 43 (KD) | 28 (KD) |
| | | 33,454 (US\$) | 70 (US\$) | 63 (US\$) | 50 (US\$) | 33 (US\$) | 143 (US\$) | 93 (US\$) |
| Store & Consumable | 1.9% | 20,325 (KD) | 42 (KD) | 38 (KD) | 29 (KD) | 19 (KD) | 87 (KD) | 57 (KD) |
| | | 67,361 (US\$) | 139 (US\$) | 126(US\$) | 96 (US\$) | 63 (US\$) | 288 (US\$) | 189 (US\$) |
| CSSD | 1.0% | 11,186 (KD) | 23 (KD) | 21 (KD) | 16 (KD) | 11 (KD) | 48 (KD) | 32 (KD) |
| | | 37,073 (US\$) | 76 (US\$) | 70 (US\$) | 53 (US\$) | 36 (US\$) | 159 (US\$) | 106 (US\$) |
| Dietetic and Food | 1.6% | 17,057 (KD) | 35 (KD) | 32 (KD) | 25 (KD) | 17 (KD) | 73 (KD) | 48 (KD) |
| | | 56,530 (US\$) | 116 (US\$) | 106 (US\$) | 83 (US\$) | 56 (US\$) | 242 (US\$) | 159 (US\$) |
| Laundry and Linen | 0.9% | 9,958 (KD) | 20 (KD) | 19 (KD) | 15 (KD) | 9 (KD) | 43 (KD) | 28 (KD) |
| | | 33,003 (US\$) | 66 (US\$) | 63 (US\$) | 50 (US\$) | 30 (US\$) | 143 (US\$) | 76 (US\$) |
| Drug | 5.5% | 58,611 (KD) | 121 (KD) | 110 (KD) | 84 (KD) | 55 (KD) | 252 (KD) | 165 (KD) |
| | | 194,249 (US\$) | 401 (US\$) | 365 (US\$) | 278 (US\$) | 182 (US\$) | 835 (US\$) | 547 (US\$) |
| Radiology | 3.7% | 40,286 (KD) | 83 (KD) | 76 (KD) | 58 (KD) | 38 (KD) | 173 (KD) | 114 (KD) |
| | | 133,516 (US\$) | 275 (US\$) | 252 (US\$) | 192 (US\$) | 126 (US\$) | 573 (US\$) | 378 (US\$) |
| Laboratory (III) | 10.2% | 109,264 (KD) | 225 (KD) | 205 (KD) | 157 (KD) | 103 (KD) | 470 (KD) | 309 (KD) |
| | | 362,123 (US\$) | 746 (US\$) | 679 (US\$) | 520 (US\$) | 341 (US\$) | 1558 (US\$) | 1,024 (US\$) |
| Physiotherapy | 4.9% | 52,791 (KD) | 109 (KD) | 99 (KD) | 76 (KD) | 50 (KD) | 227 (KD) | 149 (KD) |
| | | 174,960 (US\$) | 348 (US\$) | 328 (US\$) | 252 (US\$) | 166 (US\$) | 752 (US\$) | 494 (US\$) |
| ICU (II) | 20.6% | 221,439 (KD) | 457 (KD) | 416 (KD) | 318 (KD) | 209 (KD) | 953 (KD) | 625 (KD) |
| | | 733,893 (US\$) | 1515 (US\$) | 1379 (US\$) | 1,054 (US\$) | 693 (US\$) | 3,158 (US\$) | 2,071 (US\$) |
| Physician and Nursing (I) | 42.1% | 452,154 KD | 932 (KD) | 849 (KD) | 650 (KD) | 427 (KD) | 1,946 (KD) | 1,277 (KD) |
| | | 1,498,529 (US\$) | 3,089 (US\$) | 2,814 (US\$) | 2,154 (US\$) | 1,415 (US\$) | 6,449 (US\$) | 4,232 (US\$) |
| Average Cost | 100% | 1,074,644 (KD) | 2,216 (KD) | 2,018 (KD) | 1,544 (KD) | 1,014 (KD) | 4,626 (KD) | 3,035 (KD) |
| | | 3,561,585 (US\$) | 7,344 (US\$) | 6,688 (US\$) | 5,117 (US\$) | 3,361 (US\$) | 15,332 (US\$) | 10,059 (US\$) |

**Figure-1.**
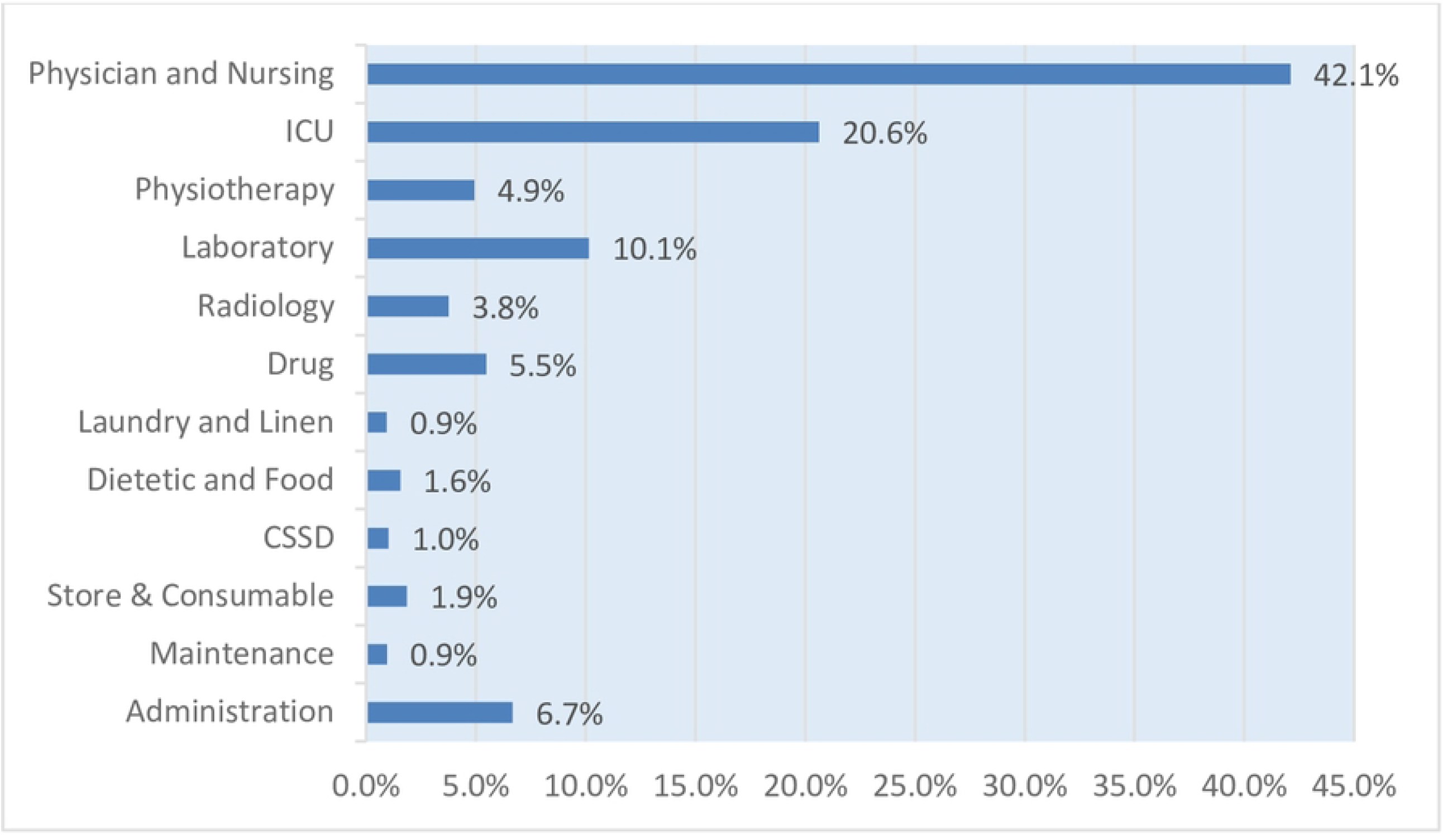
The Percentage of Cost Component in Managing Covid-19 Patients. Notes: ICU= Intensive Care Unit CSSD= Central Sterile Supply Department (CSSD)

## Discussion

This study is the first study on the cost of managing COVID-19 patients in Kuwait. The outbreak of COVID-19 and the increasing number of patients in Kuwait impose high costs on infected patients and the healthcare system. The economic burden of these diseases has been a cause of concern among managers and policymakers in the health sector. Hence, identifying the economic consequences of COVID-19 could provide valuable evidence for policymaking. This study provides an estimate of the cost of COVID-19 per patient per admission from a healthcare provider’s perspective. We report an average inpatient treatment cost of US$ 7,344, which is almost similar to the cost in China of 6,827 USD from 70 empirically observed cases [24] but is lower than the cost reported in Saudi Arabia (12, 547 USD) [25], which is also lower than that reported in a recent study conducted in the United States by Shrestha et al. (2021). They reported that the average hospitalization cost per patient was estimated to be 13,090 USD [35]. This current finding is higher compared to several studies conducted by: Bartsch et al in USA [21], where the cost of Covid-19 per ranged from 2,837 to 3,205 USD, Gedik et al in Turkey [22], where the mean cost of Covid-19 patients was 882 USD (SD=667) and Ghaffari Darab et al in Iran [23], where the mean cost of Covid-19 patients was 3,755 USD. Meanwhile, the mean cost of mild, moderate, and severe Covid-19 in this study were 3,526 USD, 6,708 USD and 15,331 USD respectively. These finding are lower compared to the study conducted by Li et al in China, the mean cost of Mild Covid-19 and moderate Covid-19 were 4,552 USD and 11,058 USD respectively [24]. In this study, it was found that more severe COVID-19 patients and ICU patients had higher healthcare costs than milder COVID-19 patients. This finding is similar to studies by Athanasakis et al [26], Gedik et al [22], Li et al [24] and Khan et al [25]. Regarding the cost comparison between the general ward and ICU admission, the treatment cost per admission in the general ward and ICU admission was US$ 5,535 and US$ 14, 390 USD, respectively. This finding is lower than the study in Saudi Arabia, where general Medical Ward (GMW) and ICU were 11,385 USD and 21,173 USD respectively [15]. This finding is also lower than that of a recent study conducted in the United States by Shrestha et al. (2021), where the average hospitalization costs per patient were estimated to be 13,090 USD without ICU and 21,222 USD with ICU admission [35]. The reason for the cost differences may be due to differences in sample size, study time period, cost methodology, and composition of sample members. There have been significant differences in the cost of healthcare delivery units worldwide in non-coronavirus circumstances [36]. Therefore, it may not be useful to compare the costs in different countries. However, such comparisons can provide an understanding of the severity of financial effects on the health systems of other countries. As shown in Table 3, the average cost of patients staying in the ICU (US$ 14,390) is about 3 times that of patient who did not receive intensive care (US$ 5,535). In the study population of the present study, 20.4% (99/485) of hospitalized patients used intensive care, while in the study by Darab et al., this rate was 7% [23]. Some other studies have suggested that about one-third of people infected with SARS-CoV-2 are in critical condition and require intensive care [37]. Others report that approximately 5% of COVID-19-confirmed cases require intensive care [38].

### Cost Component

Based on the results above (Table 4), the total treatment costs of Covid-19 inpatient were estimated to be KD 1,074,644, equal to US$ 3,561,585, in which physician and nursing care costs accounted for the largest share of costs (42.1%). The other costs were intensive care costs (20.6%; US$ 733,893), laboratory costs (10.2%; US$ 362,123), administration costs (6.7%; US$ 236,896) and drug costs (5.5%; US$ 194,249). This highest component cost is different from the Ghaffari Darab study, with the largest share of costs being intensive care and nursing services at 43% of total costs [23]. However, in the study by Li X-Z, the highest costs were related to medicines [24]. In this study, the average Length of Stay (ALOS) per patient per admission in averages: 9.4 days (SD=8.5). This ALOS is lower than a study conducted in Germany showed that although the average length of stay in hospital for all patients was 14.3 days [39]. The estimates of the present study showed that the average cost in the group with severe conditions was approximately three times that of the group with mild conditions.

The mean ± SD age of patients was estimated at 52.9 ± 18.1 years and median ages 55 years. Similar to Richardson’s study, it was also confirmed that among hospitalized patients with Covid-19, most were elderly men [40]. In a study conducted in China, hospitalized patients were mostly men, with a median age of 56 years, and 26% required inpatient care [41]. As previously mentioned, the Covid-19 outbreak has already imposed massive costs on the Kuwait healthcare system, and the disease response system must adapt to the financial shock caused by this disease in various ways. Ensuring a comprehensive response to the Covid-19 pandemic requires public funding. Reprioritizing public expenditure to strengthen the health and economic system requires timely measures by government leaders and a supportive public funding environment. To respond to these new economic and financial constraints, adjustments must be made to both the revenue and expenditure dimensions of the budget [41]. In this study, we only investigated the costs of hospitalized patients, excluding other cost dimensions in the health system and the economy of the country. Regarding the unpredictable nature of this disease and its definitive treatment, the development of evidence in the field of epidemiological dimensions and its economic effects requires appropriate decision-making and policymaking, and research efforts in this field should be continued.

## Study limitation and recommendation

This study was conducted in a single tertiary hospital assigned by the government as a center for Covid-19 patient management. The cost was calculated from the hospital perspective, excluding indirect costs such as lost income (hospitalization), lost income (recovery at home), and potential productivity lost (premature death). The cost comparison was not apple-to-apple because there were some difficulties in comparing across the literature due to differences in methodology, population, health care costs, health system, and so on. However, the study findings were analyzed using established, standardized, and reproducible methods with the aim of supporting emergency preparedness in referral hospitals in the future. This finding did not include community-based care costs, PPE equipment costs, transport costs, surveillance efforts, or other impacts on the health care system.

It is recommended that further studies look at and explore the complete cost analysis, including indirect costs such as lost income (hospitalization), lost income (recovery at home), and potential productivity loss (premature death). Other costs, such as PPE equipment cost, transport costs, out-of-pocket costs to patients, home quarantine costs, institutional quarantine costs, and vaccination costs should be included. It is suggested that the government should invest in prevention strategies rather than treatment approaches to control these diseases.

## Conclusion

The average treatment cost per patient per admission was KD 2,216 (SD=2,018), which is equal to US$ 7,344 (SD= US$ 6,688), and the average Length of Stay (ALOS) per patient per admission was 9.4 days (SD=8.5). Physician and nursing care costs accounted for the largest share of costs (42.1%; US$ 1,498,529), followed by Intensive care costs (20.6%; US$ 733,893) and laboratory costs (10.2%; US$ 362,123). The average cost for patients with severe conditions was approximately three times that of patients with mild conditions. The cost information can assist hospital managers and policymakers in designing more efficient interventions, especially for the management of high-risk groups.

## Data Availability

All relevant data are within the manuscript and its Supporting Information files. Financial information will only be available after getting approval from Hospital and finance unit of Kuwait MOH.

## Acknowledgments

We would like to acknowledge the financial support we received from Kuwait Foundation for the Advancement of Sciences (KFAS) to undertake this study. We would also like to extend our gratitude to “The Standing Committee for Coordination of Health and Medical Research, Ministry of Health, Kuwait, for providing ethical approval to conduct this research. We would also like to acknowledge Dr. Nader Al-Awadhi (Jaber Al-Ahmed Hospital Manager), Mr. Barrak Al-Hendal (Head of Cost Accounting Section Ministry of Health Kuwait), and Mrs. Shaimaa Sanaseri (Head of Medical Records Jaber Al-Ahmed Hospital) for their valuable input in this study.

## Ethical approval

The study was approved by the Research and Ethics Committee of Kuwait Ministry of Health (Approval Code:1502/2020). The study does not require informed consent because the data was extracted from Jaber Al Ahmad hospital information system database and did not involve interviews of patients.

## Data availability

The data that support the findings of this study are available from the Jaber Al-Ahmed Hospital, but restrictions apply to the availability of these data and are not publicly available. However, data are available from the authors upon reasonable request and with permission from the Jaber Al-Ahmed Hospital.

## Author Contributions

**Conceptualization**: Amrizal Muhammad Nur, Syed Mohamed Aljunid

**Data curation**: Amrizal Muhammad Nur, Mohammad Almari

**Formal analysis:** Amrizal Muhammad Nur, Syed Mohamed Aljunid

**Funding acquisition:** Amrizal Muhammad Nur, Syed Mohamed Aljunid

**Investigation:** Amrizal Muhammad Nur, Mohammad Almari

**Methodology:** Amrizal Muhammad Nur, Syed Mohamed Aljunid

**Project administration:** Amrizal Muhammad Nur and Mohammad Almari

**Resources:** Amrizal Muhammad Nur, Syed Mohamed Aljunid

**Supervision:** Amrizal Muhammad Nur, Syed Mohamed Aljunid

**Validation:** Amrizal Muhammad Nur, Syed Mohamed Aljunid

**Visualization:** Amrizal Muhammad Nur, Syed Mohamed Aljunid

**Writing – original draft:** Amrizal Muhammad Nur

**Writing – review & editing:** Amrizal Muhammad Nur, Syed Mohamed Aljunid

## References

1. Li Q, Guan X, Wu P, Wang X, Zhou L, Tong Y, et al. Early transmission dynamics in Wuhan, China, of novel coronavirus-infected pneumonia. N Engl J Med. 2020;382 (13):1199–207.

2. Chen N, Zhou M, Dong X, Qu J, Gong F, Han Y, et al. Epidemiological and clinical characteristics of 99 cases of 2019 novel coronavirus pneumonia in Wuhan, China: a descriptive study. Lancet. 2020;395(10223):507–13.

3. https://covid19.who.int/?mapFilter=cases (Accessed 30th September 2022)

4. https://covid19.who.int/region/emro/country/kw. (Accessed 30th September 2022)

5. Baker SR, Bloom N, Davis SJ, Terry SJ. COVID-Induced Economic Uncertainty. National Bureau of Economic Research Working Paper Series. 2020; No. 26983.

6. IMF. World economic outlook: The great lockdown, International Monetary Fund Report, 2020

7. Di Bidino R, Cicchetti A. Impact of SARS-CoV-2 on Provided Healthcare. Evidence From the Emergency Phase in Italy. Frontiers in Public Health. 2020;8.

8. Kringos D, Carinci F, Barbazza E, et al. Managing COVID-19 within and across health systems: why we need performance intelligence to coordinate a global response. Health Research Policy and Systems. 2020;18 (1).

9. De Oliveira Andrade R. Covid-19 is causing the collapse of Brazil’s national health service. BMJ. 2020; https://doi.org/10.1136/bmj.m3032:m3032.

10. Vernaz N, Agoritsas T, Calmy A, et al. Early experimental COVID-19 therapies: associations with length of hospital stay, mortality and related costs. Swiss Medical Weekly. 2020; https://doi.org/10.4414/smw.2020.20446.

11. Gates B. Responding to Covid-19 - A Oncein-a-Century Pandemic? N Engl J Med. 2020;382 (18):1677-9. doi: 10.1056/NEJMp2003762.

12. United Nations [Internet]. Shared responsibility, global solidarity: Responding to the socioeconomic impacts of COVID-19 2020. [Cited 12 April 2021]. Available from: https://unsdg.un.org/sites/default/files/2020-03/SG-ReportSocio-Economic-Impact-of-Covid19.pdf

13. World Health Organization [Internet]. Coronavirus disease (COVID-19) Weekly Epidemiological Update and Weekly Operational Update 2020. [Cited 11 February 2021]. Available from: https://www.who.int/emergencies/diseases/ novel-coronavirus-2019/situation-reports/

14. Lee J-W, McKibbin WJ. Estimating the global economic costs of SARS. Learning from SARS: preparing for the next disease outbreak: workshop summary. Nationa l Academies Press, Washington, DC. 2004:92.

15. AlRuthia Y, Somily AM, Alkhamali AS, Bahari OH, AlJuhani RJ, Alsenaidy M, et al. Estimation of Direct Medical Costs of Middle East Respiratory Syndrome Coronavirus Infection: A Single-Center Retrospective Chart Review Study. Infect Drug Resist. 2019; 12:3463–73. doi: 10.2147/IDR.S231087

16. World Bank [Internet]. COVID-19 (coronavirus) and the future of health financing: from resilience to sustainability 2020. [Cited 28 March 2021]. Available from: https://blogs.worldbank.org/health/covid-19-coronavirus-and-future-healthfinancing-resilience-sustainability

17. Li R, Rivers C, Tan Q, Murray MB, Toner E, Lipsitch M. Estimated Demand for US Hospital Inpatient and Intensive Care Unit Beds for Patients With COVID-19 Based on Comparisons with Wuhan and Guangzhou, China. JAMA Netw Open. 2020;3(5): e208297. doi: 10.1001/jamanetworkopen.2020.8297.

18. Shepard DS, Hodgkin D, Anthony YE, 2000. Analysis of Hospital Cost: A Manual for Managers. World Health Organization, Geneva.

19. Amrizal MN, Rohaizat Y, Zafar A, Saperi BS and Syed Aljunid (2005). Case-Mix Costing in Universiti Kebangsaan Hospital: A Top-down approach. Malaysian Journal of Public Health Medicine, 5(S2): 33–44.

20. Z Ahmed, S Aljunid, Am Nur (2010), PCV17 Estimating Cost of Inpatient Coronary Angiography Using, Stepdown Method at UKMMC, Value in Health, Vol 13, Issue 7, Pages A519.

21. Bartsch SM, Ferguson MC, McKinnell JA, et al. The potential health care costs and resource use associated with COVID-19 in the United States. Health Aff. 2020;39(6):927–35. https://doi.org/10.1377/hlthaff.2020.00426.

22. Gedik H. The cost analysis of inpatients with COVID-19. Acta Medica Mediterr. 2020;36(1):3289– 3292.

23. Ghaffari Darab M, Keshavarz K, Sadeghi E, et al. The economic burden of coronavirus disease 2019 (COVID-19): evidence from Iran. BMC Health Serv Res. 2021;21(1):132.

24. Li XZ, Jin F, Zhang J-G, et al. Treatment of coronavirus disease 2019 in Shandong, China: a cost and affordability analysis. Infect Dis Poverty. 2020;9(1):78.

25. Khan AA, AlRuthia Y, Balkhi B, et al. Survival and estimation of direct medical costs of hospitalized COVID-19 patients in the Kingdom of Saudi Arabia. Int J Environ Res Public Health. 2020;17(20):7458.

26. Athanasakis K, Nomikos N, Souliotis K, et al. PNS21 from disease burden to healthcare cost: highlighting the health economics aspects of the COVID-19 pandemic. Value Health. 2020;23: S647.

27. https://data.worldbank.org/country/KW (Accessed on 1 October 2022) (total populace dan GDP)

28. https://data.worldbank.org/indicator/SH.XPD.CHEX.GD.ZS?locations=KW (Accessed on 1 October 2022) (health expenditure)

29. https://www.exchangerates.org.uk/KWD-USD-spot-exchange-rates-history-2021.html#:~:text=This%20is%20the%20Kuwaiti%20Dinar,rate%20in%202021%3A%203.3142%20USD.

30. Jin H, Wang H, Li X, Zheng W, Ye S, Zhang S, et al. Economic burden of COVID-19, China, January March, 2020: a cost-of-illness study. Bull World Health Organ. 2021 Feb 1; 99(2):112–124. https://doi.org/10.2471/BLT.20.267112 Epub 2020 Nov 30. PMID: 33551505

31. Liang X, Xiao L, Yang X-L, Zhong X, Zhang P, Tang X, et al. Economic Burden of Public Health Care Was Higher Than That of Hospitalization and Treatment Associated With COVID-19 in China. Research Square. 2020 Sep 28 (preprint). https://doi.org/10.21203/rs.3.rs-79298/v1

32. Darab MG, Keshavarz K, Sadeghi E, Shahmohamadi J, Kavosi Z. The economic burden of coronavirus disease 2019 (COVID-19): evidence from Iran. BMC Health Serv Res. 2021 Feb 11; 21(1):132. https://doi.org/10.1186/s12913-021-06126-8 PMID: 33573650.

33. Gedik H. The cost analysis of inpatients with COVID-19. Acta Medica Mediterr. 2020; 36(6):3289. https://doi.org/10.19193/0393-6384_2020_6_520

34. Miethke-Morais A, Cassenote A, Piva H, Tokunaga E, Cobello V, Rodrigues Gonçalves FA, et al. Unraveling COVID-19-related hospital costs: The impact of clinical and demographic conditions. medRxiv. 2020 Jan 1;2020.12.24.20248633. https://doi.org/10.1101/2020.12.24.20248633

35. Sundar S. Shrestha, Lyudmyla Kompaniyets, Scott D. Grosse, Aaron M. Harris, James Baggs, Kanta Sircar and Adi V. Gundlapalli, Estimation of Coronavirus Disease 2019 Hospitalization Costs from a Large Electronic Administrative Discharge Database, March 2020–July 2021. Open Froum Infectious Diseases, 2021: 1–7

36. Moses MW, Pedroza P, Baral R, Bloom S, Brown J, Chapin A, et al. Funding and services needed to achieve universal health coverage: applications of global, regional, and national estimates of utilisation of outpatient visits and inpatient admissions from 1990 to 2016, and unit costs from 1995 to 2016. Lancet Public Health. 2019;4(1):e49–e73. doi: 10.1016/S2468-2667(18)30213-5.

37. Liew MF, Siow WT, MacLaren G, See KC. Preparing for COVID-19: early experience from an intensive care unit in Singapore. Crit Care. 2020;24(1):83. doi: 10.1186/s13054-020-2814-x.

38. Murthy S, Gomersall CD, Fowler RA. Care for Critically Ill Patients With COVID-19. JAMA. 2020;323(15):1499–500. doi: 10.1001/jama.2020.3633.

39. Karagiannidis C, Mostert C, Hentschker C, Voshaar T, Malzahn J, Schillinger G, et al. Case characteristics, resource use, and outcomes of 10 021 patients with COVID-19 admitted to 920 German hospitals: an observational study. Lancet Respir Med. 2020;8(9):853–62. doi: 10.1016/S2213-2600(20)30316-7

40. Richardson S, Hirsch JS, Narasimhan M, Crawford JM, McGinn T, Davidson KW, et al. Presenting Characteristics, Comorbidities, and Outcomes Among 5700 Patients Hospitalized With COVID-19 in the New York City Area. JAMA. 2020;323(20):2052–9. doi: 10.1001/jama.2020.6775

41. Zhou F, Yu T, Du R, Fan G, Liu Y, Liu Z, et al. Clinical course and risk factors for mortality of adult inpatients with COVID-19 in Wuhan, China: a retrospective cohort study. Lancet. 2020;395(10229):1054-

